# Using Explainable AI to understand frailty indicators

**DOI:** 10.1101/2023.05.30.23290760

**Authors:** Attayeb Mohsen, Masaki Yamamoto, Agustin Martin-Morales, Daiki Watanabe, Nobuo Nishi, Takashi Nakagata, Tsukasa Yoshida, Motohiko Miyachi, Kenji Mizuguchi, Michihiro Araki

**Author notes:** Corresponding authors: Email addresses (Attayeb Mohsen), (Motohiko Miyachi), (Michihiro Araki).

## Abstract

**Introduction:** The prevalence of frailty is on the rise with the aging population and increasing life expectancy, which often is accompanied by comorbidities. Frailty can be effectively detected using Frailty index such as KCL index. Early detection of frailty allows applying measures that reduce the conversion rate to frail, and improve the quality of life in the frail people. Therefore, to facilitate the screening of frailty status at the primary care level, we suggest to produce a shorter version of the KCL questionnaire.

**Aim:** To understand the importance of KCL components in the decision making process for frailty and use machine learning approach to shorten the Questionnaire while maintaining reasonable accuracy, making it easier to screen for frailty in primary care.

**Methods:** We developed an automated framework of three steps: Feature importance determination using Shap values, testing models with Cross-validation with increased addition of selected features. Moreover, we validated the reliability of KCL to detect frailty by comparing the results of KCL criteria with the unsupervised clustering of the data.

**Results:** Our approach allowed us to identify the most important questions in the KCL questionnaire and demonstrate its performance using a short version with only four questions (4) Do you visit homes of friends?, (6) Are you able to go upstairs without using handrails or the wall for support? (10) Do you feel anxious about falling when you walk?, and (25) (In the past two weeks) Have you felt exhausted for no apparent reason?). We also showed that the data clustering corresponds well with the results of KCL criteria.

**Discussion and Conclusion:** While it is difficult to predict pre-frail status using shorter KCL questionnaire, it was shown to be fairly accurate in predicting frail status using only four questions.

## 1. Introduction

A frail person can be described as a person who is barely doing well, which implies that the person will break and his health will collapse as soon as they are exposed to even minor stress or an accident, such as falling or mild infection. Therefore, frailty can be defined as the antonym of fitness/fit[1, 2]. The course of frailty has been described as “a decline in functioning across multiple physiological systems, accompanied by an increased vulnerability to stressors”[3]. Studies have also shown that frail individuals are more likely to experience falls, hospitalization, and death[4, 5], which can greatly impact their quality of life and put a strain on the healthcare system and economy[6].

It is possible for frail people to transit to robust or the other way around. That was corroborated by Japan’s geriatric society when they changed their understanding of frailty from the natural decline of a person’s fitness, health, and psychological well-being, to a preventable condition that needs attention and cure in 2014[7].

Diagnosis of frailty should start at the primary care level, where the patient is typically treated comprehensively. The early assessment of frailty makes early detection of frailty status possible, which subsequently help prevent conversion of robust adults to frail, as well as facilitate the returning from frail to robust[8] by applying appropriate preventive and treatment measures. Over five years of follow-up, adults with low body mass index and more physical activity are more likely to remain robust than the others, suggesting that preventing conversion to frail by modifying their exercise behavior and dietary habits is possible [9].

Frailty assessment is built on two main models: (1) the phenotype model and its evaluation method is Cardiovascular Health Study (CHS) criteria created by Fried et al. [4], and (2) the accumulation deficit model and its representative evaluation method, the Fraily Index (FI), advocated by Rockwood et al. [10].

Fried et al. built their Cardiovascular Health Study (CHS) Criteria on the presence of three out of these five components: Shrinkage (weight loss), Weakness, Poor endurance, slowness, and low physical activity level [4].

The Asia-Pacific Clinical Practice Guidelines for Management of Frailty evaluated multiple frailty measurements and classified them based on their use in clinical practice. The Frailty Index (FI) and Kihon Checklist were both classified as assessment tools that not only assess physical domains, but also comprehensively include mental or psychological domains. [2]. A good frailty index can be created by counting the number of symptoms or signs a subject has, the frailty status of that subject correlates with that number. Moreover, the frailty index is consistent even if the symptoms or signs are not the same[11].

The Kihon Checklist (KCL) system [12, 13], that consists of 25-items, was developed and published by Japanese Ministry of Health, Labour and Welfare (MHLW) in 2006[14, 15]. Although KCL was developed initially to predict if individual requires long-term care, is has been used extensively as an evaluation of frailty due to its similarity to FI and its comparable results with CHS[16].

Recent research extensively evaluated the KCL system, and found it useful in predicting the need for long-term care, and the mortality risk. It also shows that KCL has higher accuracy among other frailty tools[17]. It revealed that the cutoff value of 7 points or more of 25 to define frailty is appropriate[16, 18, 19]. Currently, KCL is globally used and translated into various languages[12, 20–24].

Previous studies have employed machine learning to predict frailty using data of specific questionnaire with other measurements such as handgrip strength and walking speed[25]. They suggest that their models is particularly important in detecting the frailty in early stages, to facilitate prevention of further deterioration. Hassler et al. built predictive models for frailty and applied feature selection using the Boruta algorithm. They also examined the model performance by applying a 10-fold cross-validation approach. The dataset analyzed here has more than 200 variables. They fused the class pre-frail and frail into one class. Furthermore, they built a binary classification model. Because the amount of missed data was significant in some variables, they needed to take many steps to preprocess the data, including imputing the missed values [26]. Aponte-Hao et al. examined the usage of machine learning to predict frailty in Canadian primary care clinics. They used a hold-out test set and examined eight supervised machine-learning algorithms. Their data included other features than our paper, such as diagnosis, billing codes, and medicine prescribed. Aponte-Hao et al. used a simple feature selection approach by removing the variables with less variation. Moreover, they removed the data with a high prevalence of missing values. The most important features were age, sex, and previous medical diagnosis. The sensitivity and specificity of the produced models were topped at 80% and 77%, respectively. [27]. The important point of these models is that they use regularly collected data in primary clinical care visits to predict frailty status. Kuo et al. built a machine-learning model to predict social frailty. Interestingly, they showed that one of the crucial predictors is religious participation (number 3) following health literacy and comorbidity. Religious participation item in here is similar to the KCL question about visiting a friend’s house. They both reflect social and psychological well-being as well as musculoskeletal health [28]. Tseng et al. studied the prediction of cognitive frailty among the old Taiwanese cohort. Using logistic regression, they identified six factors associated with frailty, most related to age, gender and body weight, memory deficit, and medical history of diabetes mellitus [29].

More efforts are required to assess frailty and pull attention towards prevention and treatment[7]. New, more accessible approaches are required to diagnose frailty at the primary care level. Previously, Searle et al. suggested that using a high number of items in the frailty index leads to a more accurate diagnosis. However, this approach is built on the assumption that all symptoms are equally important on the assessment of frailty, while in reality their impact is sometimes different. In this study we will try a different approach, which is to use explainable AI to understand the importance of the frailty index items using the data (data-driven approach).

This study aims to prioritize the questions in KCL according to their importance and contribution to the decision-making process. and subsequently, create a shorter version of KCL and a more straightforward interpretation using Explainable AI.

## 2. Methods

### 2.1. Subjects

The subjects of this study were randomly selected among people of 40 years or older in Settsu (February –March 2019) and Hannan (January –February 2020) cities from Osaka prefecture, Japan. The participants were stratified by Age and Sex, and they were evenly distributed geographically using school zones[30, 31]. The study questionnaire was submitted by mail. The total number of post-mails sent was 10000 in Settsu city and 8000 in Hannan city. The number of participants was 5809 (58 %) and 4801 (60 %), respectively. Later on, the responses with incomplete answers were excluded, and the total number went for the subsequent analysis steps was 7400 responses. The age and gender of the participants is summarized in table 1. Scores of less than 4 were considered robust, from 4 to 6 were considered pre-frail, and scores of 7 or more were considered frail.

**Table 1:**
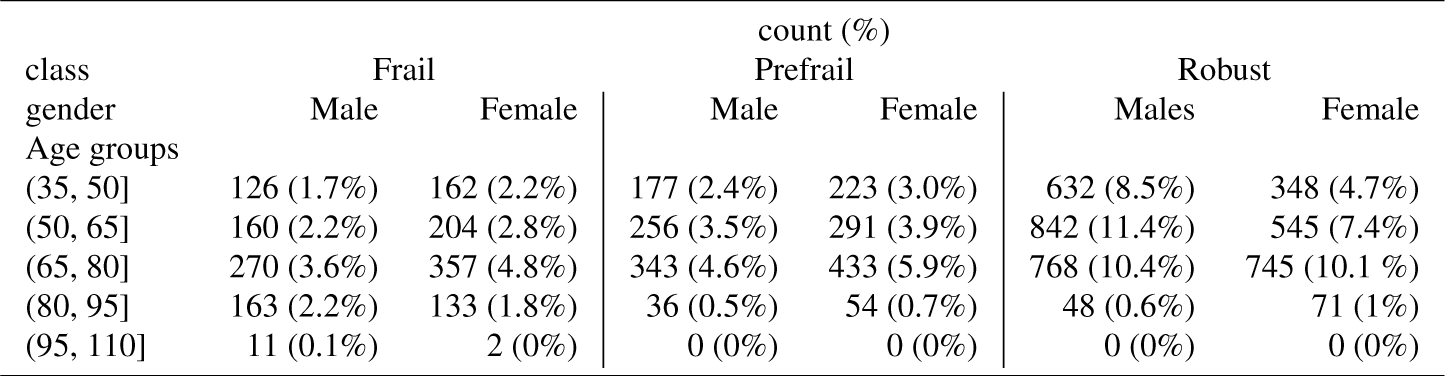
Age, gender and frailty status of participants.

The data analyzed in this paper was the KCL questionnaire and Food intake frequency survey.

### 2.2. Machine learning feature selection

To gain insight into how individual KCL questionnaire question informs the classification process of subjects, we followed a novel approach of combining machine learning models building with cross validation evaluation, then feature selection and models building again using those features in order. the code of this approach is available in https://github.com/attayeb/proml.

We built multiple machine learning models (Random forest, Gradient boost, SVM, Logistic regression, Decision tree, and Gaussian Naive Bayes) and validatated them. First, shapely values were calculated for each model, and the top features were identified. Then, multiple models are created for each machine learning algorithm using the top two features at the beginning, with ten folds of cross-validations. Next, the average of Matthews correlation coefficients was calculated for all the cross-validation folds. Then progressively adding one feature at a time, repeat the same steps. Finally, the top-performing models were visualized to show how features contribute to the accuracy of the prediction models (Check Algorithm 1). The data used for building the models were all the data set, with all variables. Three models were built, to differentiate between all pairs of subject frailty classes. The default parameters according to sci-kit learn package were used. We chose Matthews correlation coefficient because it is recommended in imbalanced data sets [32].

**Algorithm 1:**
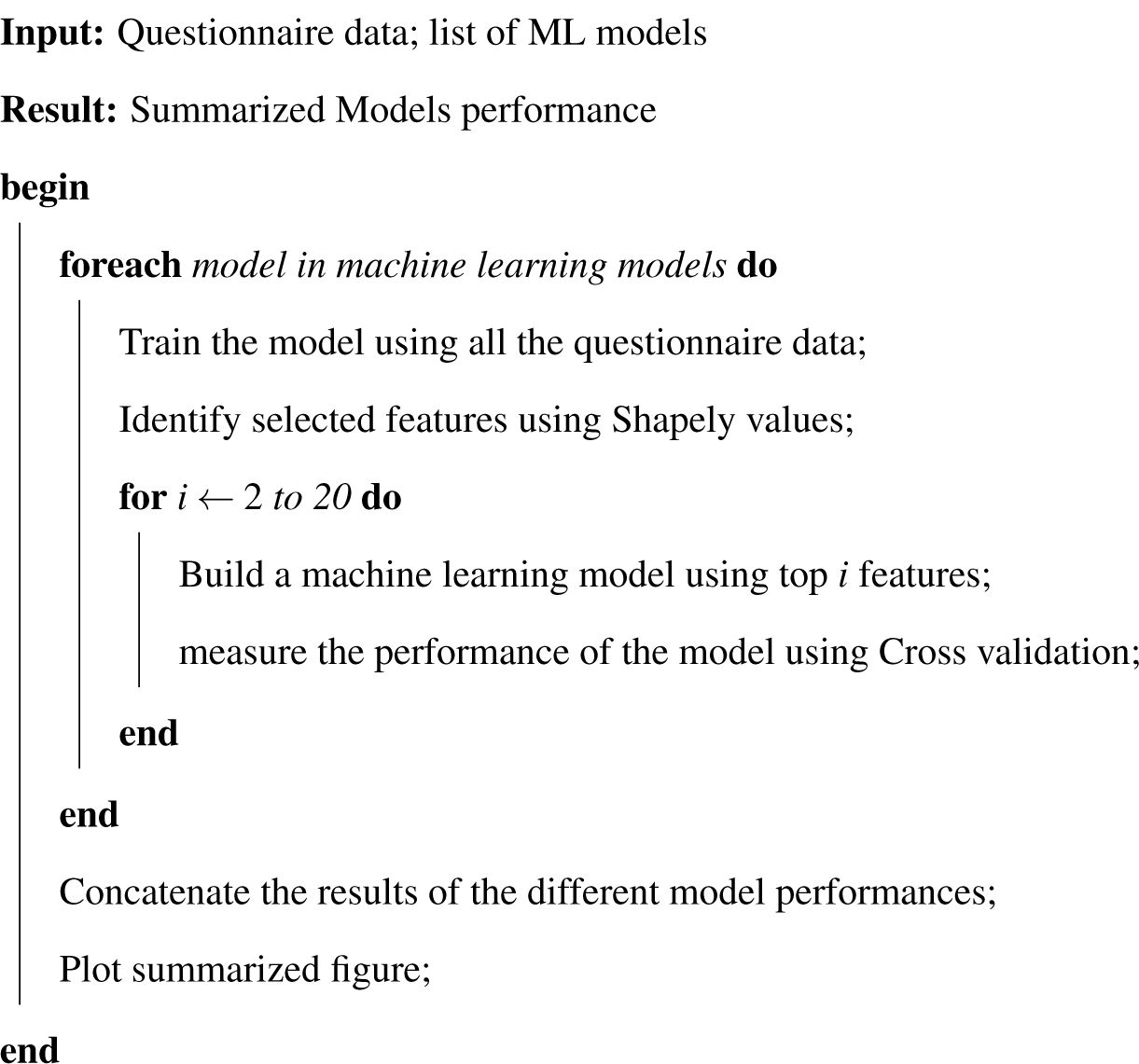
Machine learning models evaluation

### 2.3. Subgroup detection

The original classification of subjects was determined based on the KCL questionnaire criteria, which utilizes the same data we used to build our classification models and feature selection process. This requires further evaluation if the data is possibly clusterable to clusters that corresponds at least partially with the criteria results. To do that, we examined if the questionnaire results could be clustered using unsupervised clustering algorithms. We reduced the data dimensions to 2 dimensions and plotted them. We tried PCA (Prinicipal component analysis) [33], t-SNE (t-Distributed Stochastic Neighbor Embedding) [34] and UMAP (Uniform Manifold Approximation and Projection for Dimension Reduction) [35]. Before running these algorithms dimension reduction, we applied quantile transformation for data scaling using scikit-learn [36] QuantileTransformer function.

### 2.4. Used packages

All model training, validation, and feature selection was done in local system using Ubuntu 20.04. All the analysis was done in Python programming language version 3.9; For PCA, RandomForest, SVM, and logistic regression, we used scikit-learn package [36], for feature importance, we euse SHAP (SHapley Additive exPlanations) package [37].

## 3. Results

### 3.1. Progressive feature contribution assessment

To understand the impact of the questions on the classification accuracy, we applied shapely values algorithm to prioritize these questions depending on their contribution to the decision of the prediction model, then we examined the performance of the model by adding the features one by one progressively starting with the top features with the greatest importance. This approach demonstrates the importance of any question in the final classification of the case. Addition of one question at a time make it possible to decide at what number of questions the accuracy of the prediction using machine learning is enough.

Classifying subject into either Robust or Frail does not require many questions to reach reasonable performance measured by Matthews’ correlation coefficient (Figure 1). Only four questions were required: namely, “Do you visit home of friends?”, “Are you able to go upstairs without using handrails or the wall for support?”, “Do you feel anxious about falling when you walk?” and “In the past two weeks, have you felt exhausted in no apparent reasons?” can successfully differentiate Robust from frail classes. The accuracy using these 4 questions with Gradient boost model with 0.725 of Matthews correlation coefficient values.

**Figure 1:**
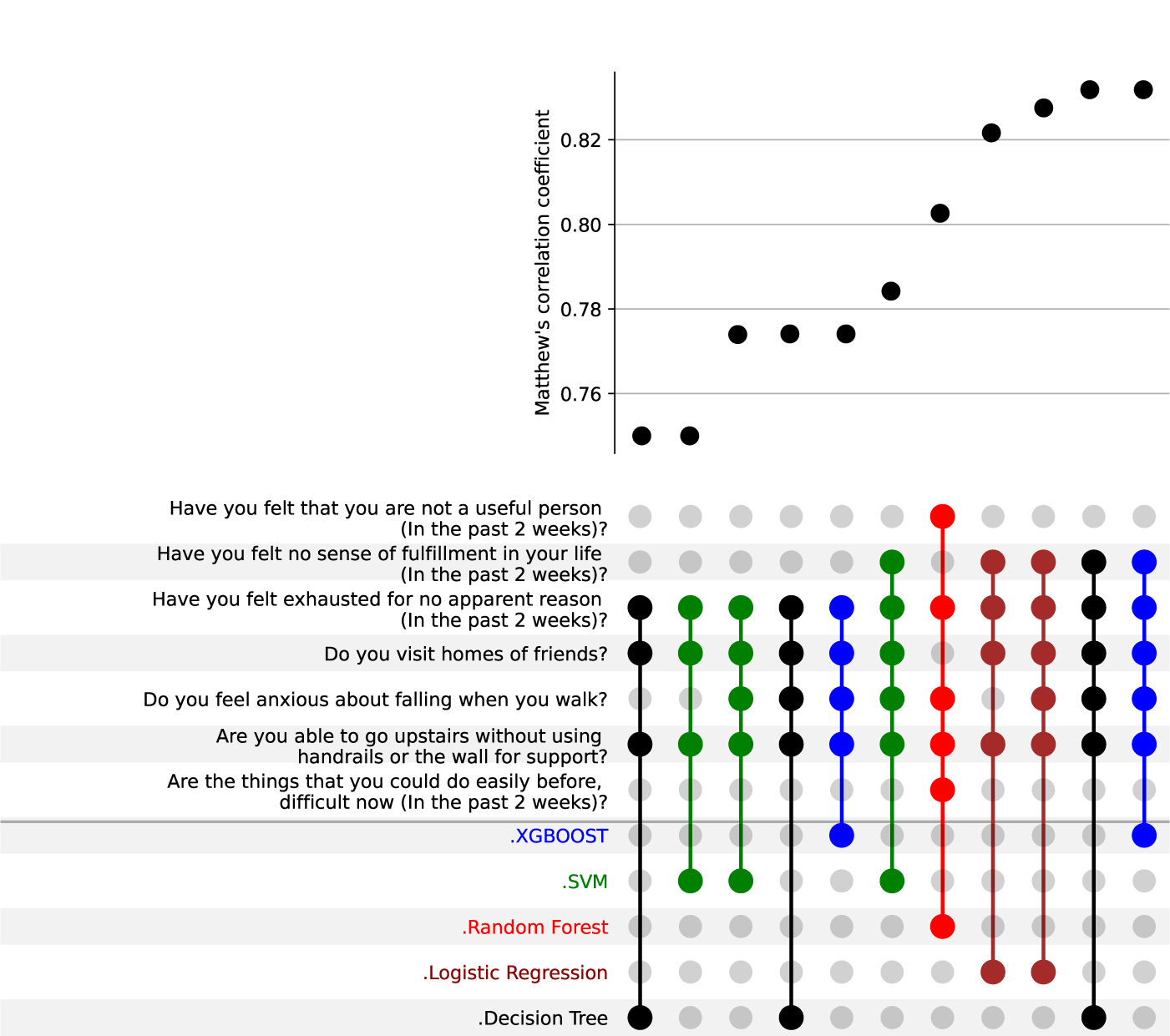
Progressive feature contribution assessment for Robust vs Frail. Small number of features are required to differentiate (as little as 4 questions can give prediction with 0.75 of Matthews correlation coefficient).

Those four questions were the most commonly selected over Decision Tree, Gradient Boost, and logistic regression.

A higher number of questions is required to accurately identify subjects as Robust or Pre-frail (as shown in Figure 2). Around ten questions are required to have MCC values of more than 0.6; those questions were shared among machine learning algorithms demonstrating the consistency of their relevance to the decision process.

**Figure 2:**
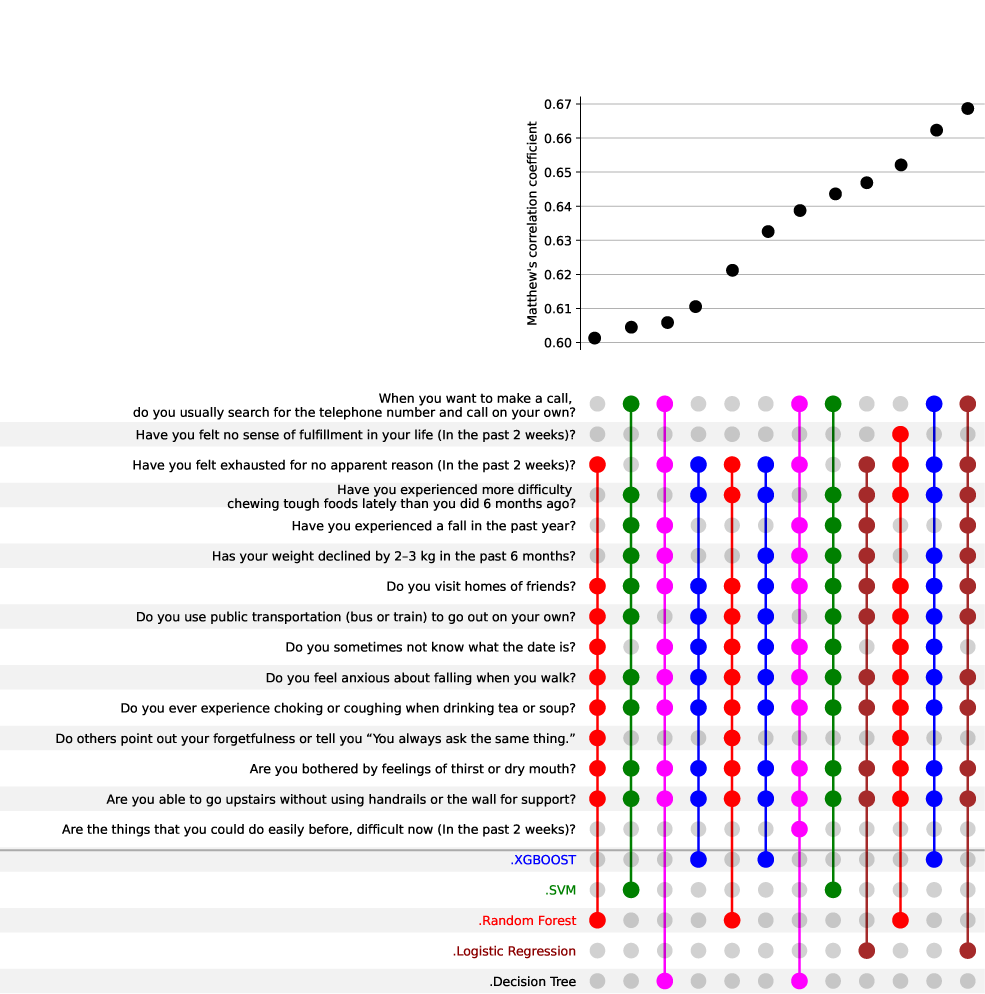
Progressive feature contribution assessment in differentiating Robust from Pre-frail classes, color represents the machine learning modalities and the questions was added gradually to find the best combination of questions that lead to better prediction. (MCC: Matthews Correlation Coefficient).

Frail with Pre-frail is also a challenging task. It can be differentiated by asking a higher number of questions, including “Do you use public transportation”, “Are you bothered by the feeling of thirst or dry mouth?” and “In the past two weeks, are the things that you could do easily before, difficult now?”. The accuracy was around more than 0.65 Matthews correlation coefficient.

### 3.2. Suggested short Questionnaire

Creating a shorter version of KCL questionnaire can help increase the usability of KCL questionnaire, having less number of question makes the screening less time consuming and more straight forward. To do that, we chose the top 4 crucial questions identified in the Frail-Robust model, we built Random Forest model. We created a sham dataset with all possible combination of answers (16) and then calculated the probability of classification as robust (Check Table 2).

**Table 2:** Short version of Kihon Questionnaire. (✓): answering Yes, (✗): answering No. Four questions are selected, the “Robustness probabiliry” columns shows the probability of being robust using the answers of these 4 questions.

| Do you visit homes of friends? | Are you able to go upstairs without using handrails or the wall for support? | Do you feel anxious about falling when you walk? | (In the past 2 weeks) Have you felt exhausted for no apparent reason? | Robustness Probability |
| --- | --- | --- | --- | --- |
| ✓ | ✓ | ✓ | ✓ | 72 |
| ✓ | ✓ | ✓ | ✗ | 99 |
| ✓ | ✓ | ✗ | ✓ | 99 |
| ✓ | ✓ | ✗ | ✗ | 100 |
| ✓ | ✗ | ✓ | ✓ | 16 |
| ✓ | ✗ | ✓ | ✗ | 86 |
| ✓ | ✗ | ✗ | ✓ | 54 |
| ✓ | ✗ | ✗ | ✗ | 100 |
| ✗ | ✓ | ✓ | ✓ | 12 |
| ✗ | ✓ | ✓ | ✗ | 79 |
| ✗ | ✓ | ✗ | ✓ | 50 |
| ✗ | ✓ | ✗ | ✗ | 97 |
| ✗ | ✗ | ✓ | ✓ | 0 |
| ✗ | ✗ | ✓ | ✗ | 30 |
| ✗ | ✗ | ✗ | ✓ | 8 |
| ✗ | ✗ | ✗ | ✗ | 75 |

### 3.3. Subgroup discovery

To further reinforce our conclusion of the selected questions, we showed the data items collected can be clustered to groups corresponds to the KCL criteria classificatin (Figures 4, 5, 6). The segregation is more prominent when it comes to Frail compared to the pre-frail and Robust classes, however, the separation between Pre-frail and Robust was less obvious (Figures 4C, 5C, 6C).

**Figure 3:**
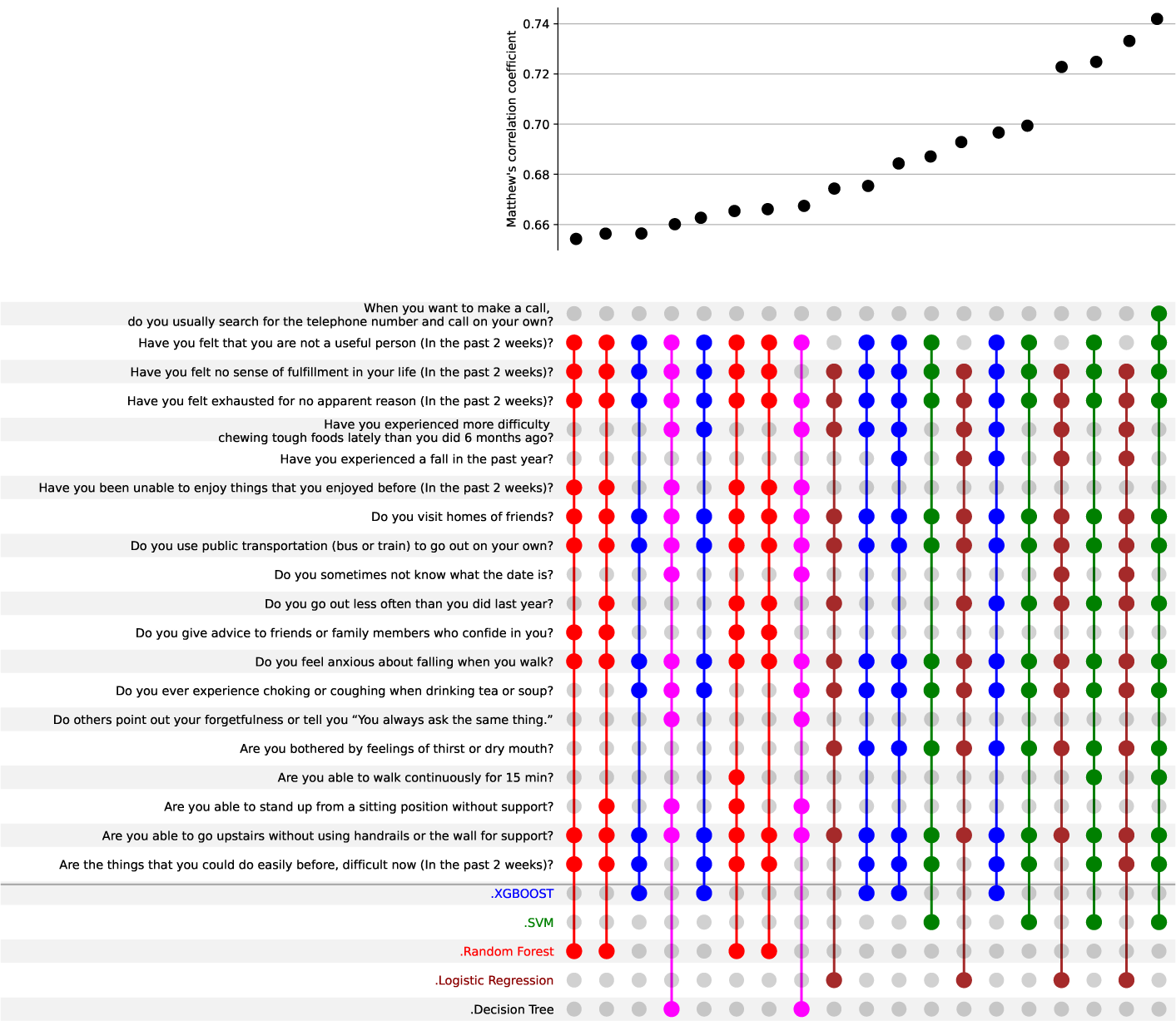
Progressive feature contribution assessment in differentiating frail from Pre-frail classes, color represents the machine learning modalities and the questions were added gradually to find the best combination of questions that lead to better prediction. (MCC: Matthews Correlation Coefficient).

**Figure 4:**
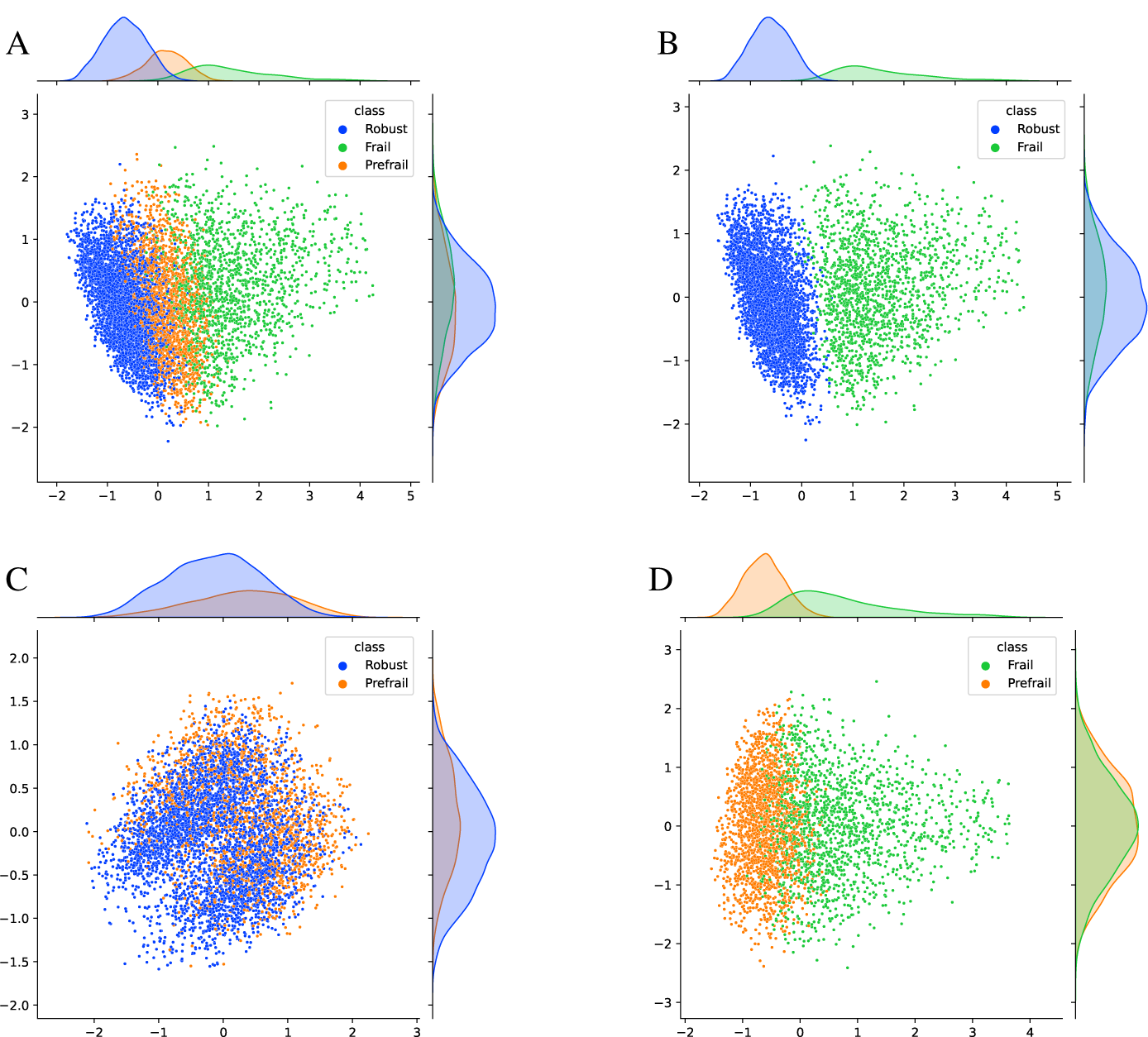
Distribution of subjects in two dimensional PCA (principle component analysis) of questionnaire answers regarding to their class. **A.** all three classes, **B.** Robust and Frail classes, C. Robust and Prefrail, **D.** Frail and prefrail. Principle component analysis was measured with 2 components. A, B, and D showed good subgroup discovery in contract to C. (top and right side shows density plot of first and second components respectively)

**Figure 5:**
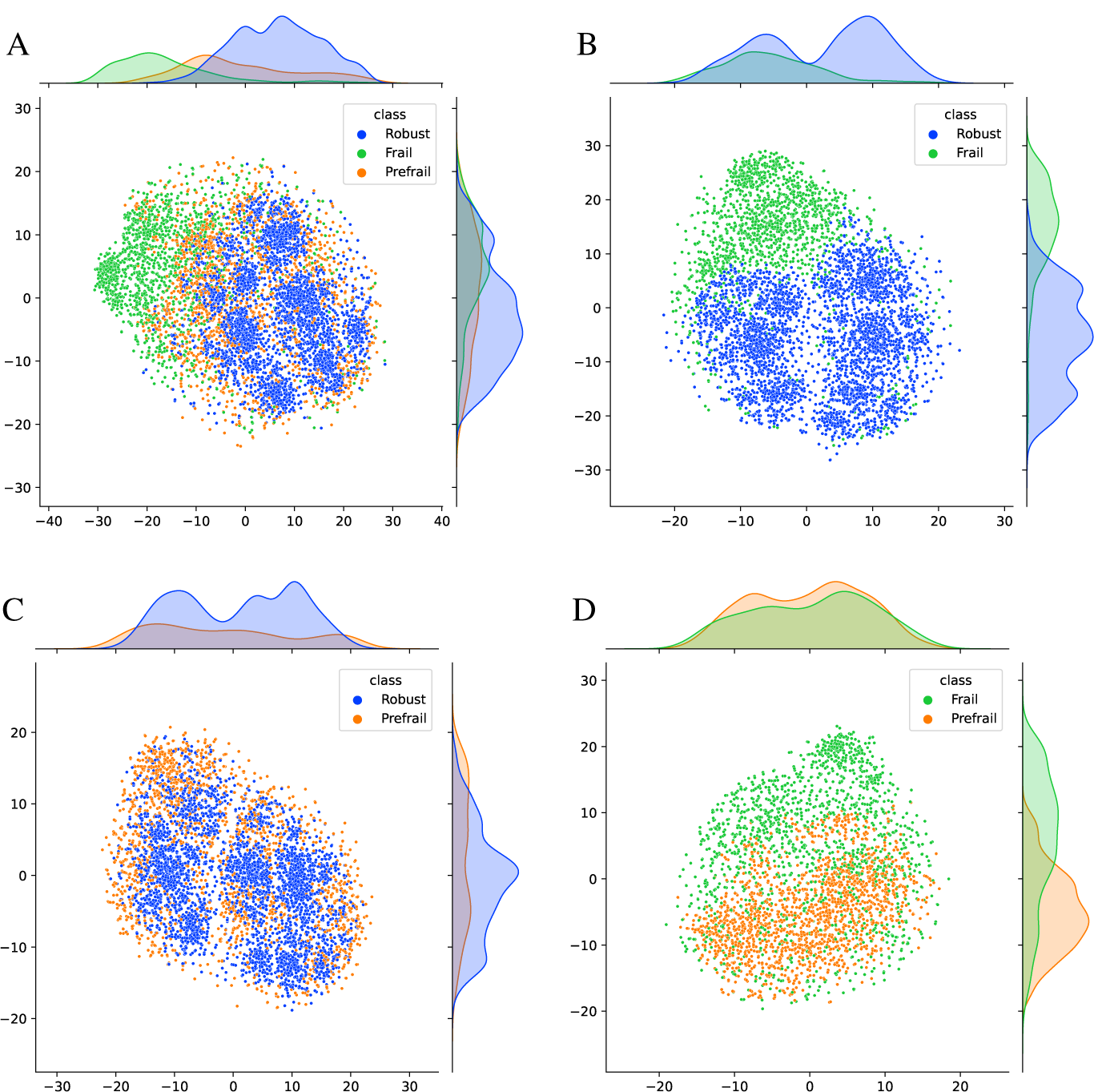
Distribution of subjects in two dimensional t-Distributed Stochastic Neighbor Embedding (t-SNE) of questionnaire answers regarding to their class. **A.** all three classes, **B.** Robust and Frail classes, C. Robust and Prefrail, **D.** Frail and prefrail. t-SNE embedding was plotted in 2 components. A, B, and D showed good subgroup discovery in contract to C. (top and right side shows density plot of first and second dimensions respectively)

**Figure 6:**
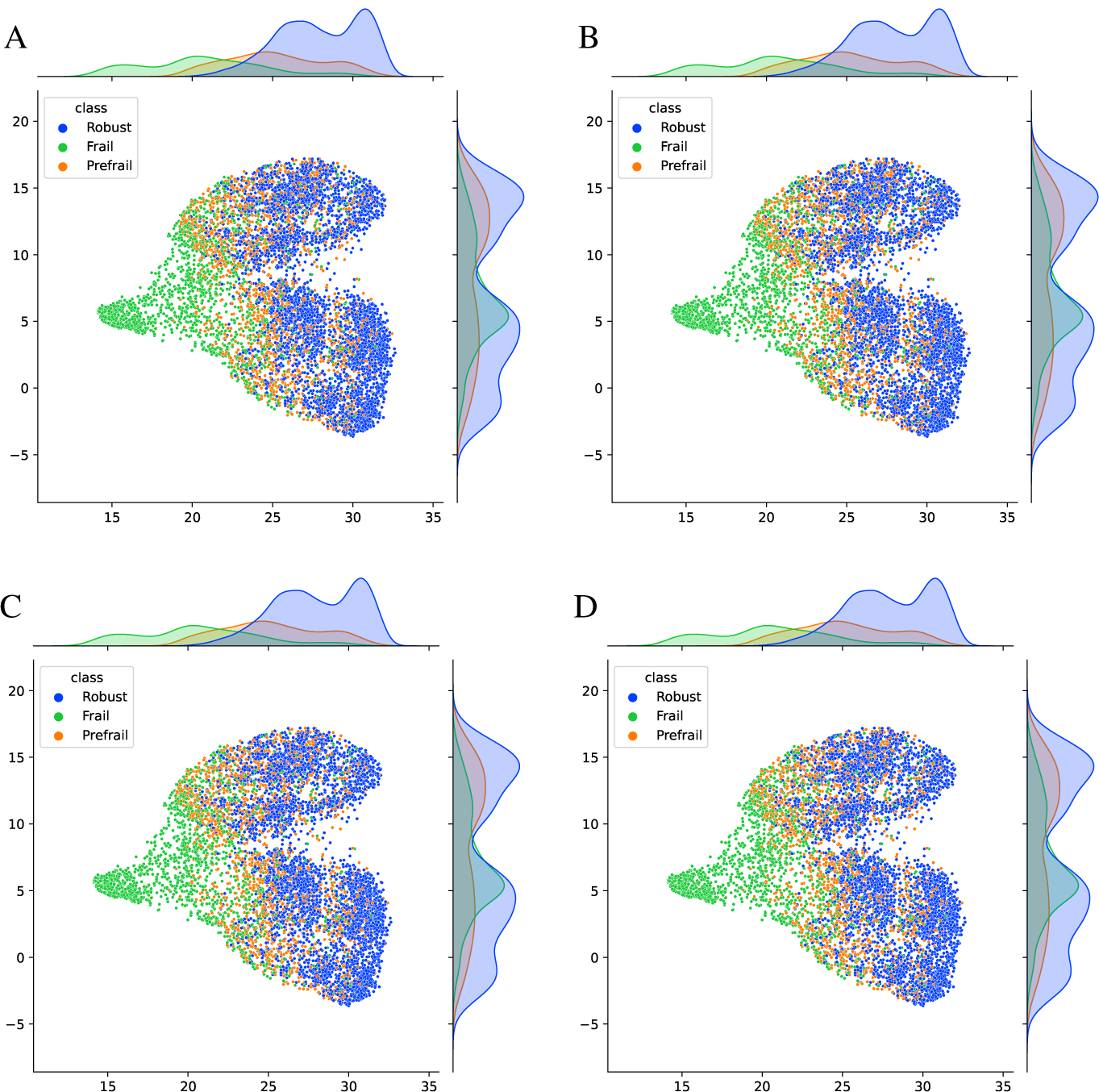
Distribution of subjects in two dimensional Uniform Manifold Approximation and Projection (UMAP) of questionnaire answers regarding to their class. **A.** all three classes, **B.** Robust and Frail classes, C. Robust and Prefrail, **D.** Frail and prefrail. UMAP was measured and plotted in 2 dimensions. A, B, and D showed good subgroup discovery in contract to C. (top and right side shows density plot of first and second dimensions respectively)

### 3.4. Class-cluster overlap

To examine if the questionnaire can correctly classify the data using unsupervised clustering method and at the same time understand how robust is that, we clustered the data to three clusters using Kmeans clustering algorithm. The close examination of the resulting clusters (Figure 7) shows that majority of cluster 3 belongs to Robust class, and the majority of cluster 1 belongs to Frail class, however, the Pre-frail group is divided between the three clusters, and cluster2 contained mainly subjects from Robust and Pre-frail. This observation suggests that the questionnaire robustness was higher when it comes to differentiate Robust from frail, however determination of Pre-frail class is more difficult.

**Figure 7:**
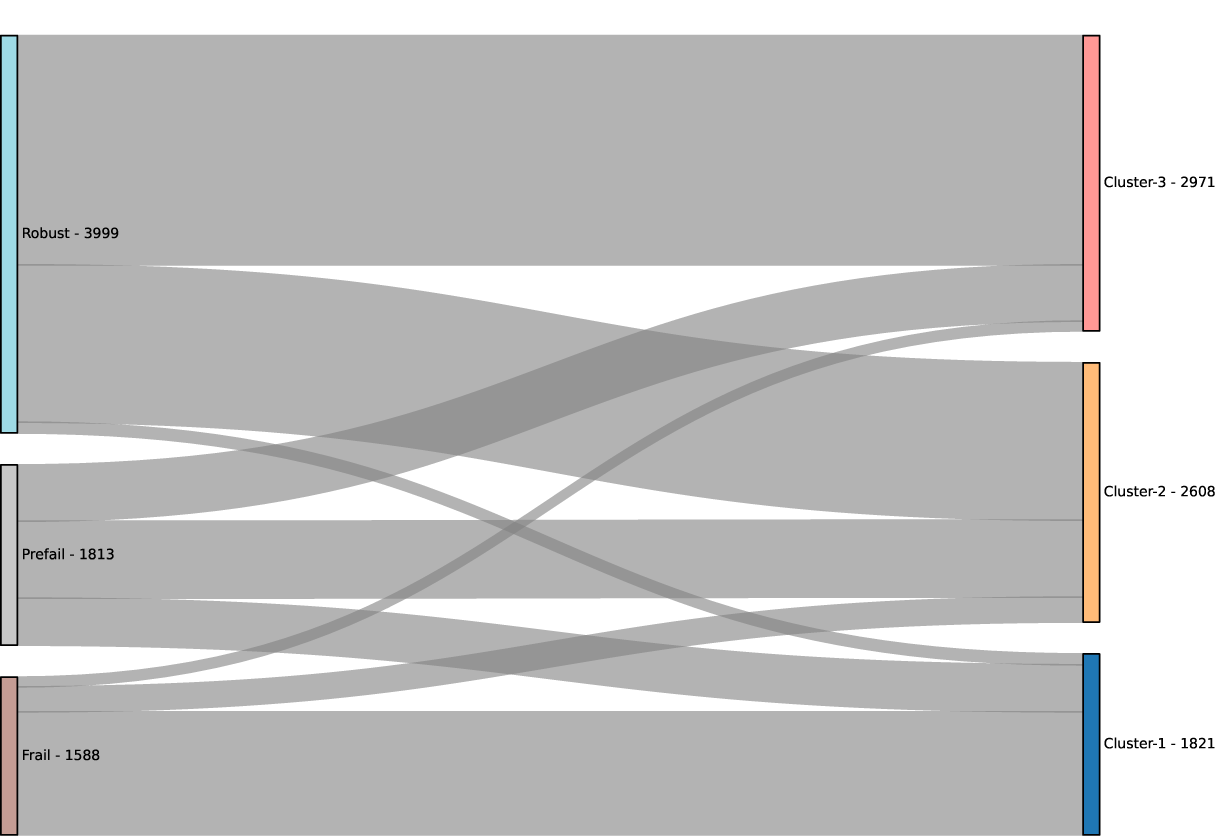
Kmeans clustering compatibility with the original frailty classes. Most of robust subjects belong to cluster 3, and Frail to cluster 1, however, prefrail was not clearly clustered in one cluster, it was dispersed between the three clusters.

To provide a more comprehensive understanding of sample clustering and to compare the resulting clusters with the original classification, we re-analyzed the data using the Kmeans algorithm with 10 clusters. This approach allowed for a more flexible and informative depiction compared to the initial clustering of three clusters. Additionally, we calculated the overlap between the resulting clusters and the original classes (Figure 8). There are no substantially shared clusters between Robust and Frail, i.e. any given cluster should belong to either one of them but not both. but when it comes to pre-frail, There is no single cluster belongs to Pre-frail with 0.5 or more overlap. They are always shared between pre-frail and robust, or pre-frail and Frail.

**Figure 8:**
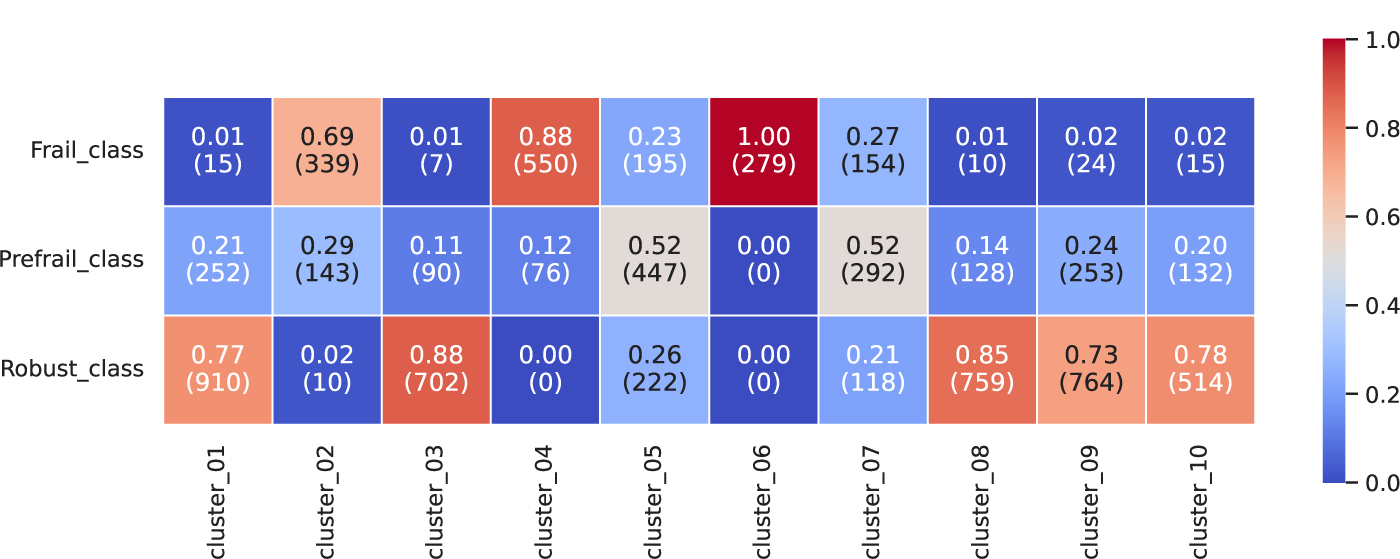
Overlap heatmap between three classes and 10 data clusters. Clusters [1, 3, 8, 9 and 10] have higher overlap with Robust class, meanwhile clusters [2, 4, 6] overlaps more with frail class. Half of clusters [5, 7] members belong to prefrail class, and the other half is equally distributed between the other two classes.

In Figure 9, we clustered the data points to two clusters, plot them after reducing the dimensions using factor analysis. Most of Robust and Prefrail were clustered in one cluster, and frail was clustered alone. We calculated Matthews correlation coefficient after merging Prefrail and Robust, the clusters shows a value of 0.835, which suggested very high similarity between the original classes of data and the clusters.

**Figure 9:**
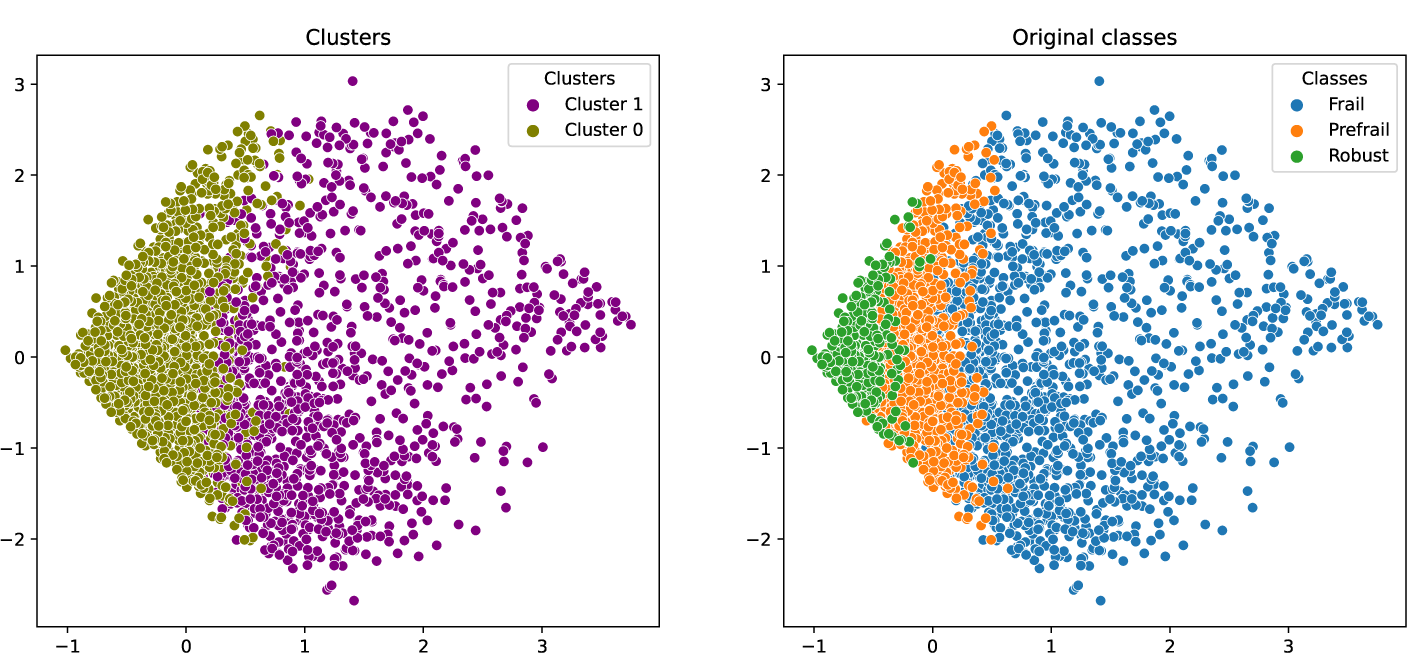
Clustering the data to two clusters using KMeans, and presenting them in two dimensions after Factor analysis dimension reduction. Left side shows the clusters, and the right side shows the original classes identified by KCL criteria.

These results suggested that the questions in the questionnaire was best in differentiating Frail from Robust, and at the same time, they could separate between Pre-frail and the other two classes with less accuracy.

## 4. Discussion

In this paper, we ranked the KCL questions by their significance in determining frailty status. We also expanded these results by proposing a simplified KCL questionnaire that can be used for screening purposes through a novel approach that integrates feature selection and the gradual construction and evaluation of machine learning models. We also validated the usability of the data collected using the KCL questionnaire by unsupervised clustering that produced two distinctive clusters (Frail and Robust) and the third is an intermediate cluster (Pre-frail). The ability of KCL to produce these three clusters suggests the robustness of KCL as an indicator of frailty.

The responders of this questionnaire were assigned to three groups of robust, frail and pre-frail following simple criteria that considers the number of questions answered. To validate if this survey is sensitive, we cluster the subjects’ responses using multiple unsupervised methods. The objective of this clustering is to confirm if the data can be adequately classified without any intervention. Figures (4, 5, 6, 7, and 8) demonstrate the clear separation between the clusters of frail and robust subjects, with the prefrail class occupying the intermediate position. That is more prominent in figure 8, where no cluster has a similar number of individuals shared among the three groups or particularly among frail and robust. This observation implies that the questionnaire was designed and interpreted in a manner that produces reliable results. Had some questions were not carefully chosen or not being informative, the resulting clusters would contain a cluster or more with majority of both robust and frail.

The early diagnosis of frailty paves the road to recovery and the patient’s protection against quick deterioration. Therefore, creating a frailty test that can be easily used for screening is crucial [8].

Frailty research has extensively utilized machine learning [25–29] and explainable AI [38] That can shed light on the mechanisms of diseases and identify biomarkers that can be used to diagnose or find a treatment for a specific medical condition. Moreover, it can be extended to other applications, such as understanding the prognostic factors or severity modulators[39].

Using machine learning and explainable AI allowed us to produce a shorter version of the KCL questionnaire with reasonable accuracy. Moreover, The questions selected (Table: 2) reflect both physical and psychological status. For instance, the question “(4) Do you visit homes of friends?” reflects the social well-being of the subject, which includes psychological status but it also extends to motor activities. Conversely, the subject’s inability or disinterest in visiting friends’ homes could stem from either or both physical and psychological limitations. The second question about going upstairs without using handrails or the wall for support is mainly a physical indicator. However, if the subject has safety concerns or doubts about their abilities, they may still use handrails, thus also serving as a psychological indicator. This relation between physical and mental health is further evident in the question “(10) Do you feel anxious about falling when you walk?” which similarly reflects both aspects of the subject’s health.

The data showed in this paper suggest that using a combination of comprehensive questions showed a better indication of frailty than purely physical or psychological questions such as “Are you able to stand up from a sitting position without support?” or “Are you able to walk continuously for 15 min?”.

The accuracy of the result simplified questionnaire is lower than the accuracy if we use the entire questionnaire. Also, our method showed it is difficult to predict the pre-frail state using a smaller number of KCL questions. This limitation arises from the fact of resemblance between pre-frail and Robust.

## 5. Conclusions

In this paper we examined the performance of KCL questionnaire using a novel machine learning-feature selection approach. We identified and prioritized the questions according to their contribution in the classification decision made. Moreover, we suggest a shortened version of KCL questionnaire with 4 questions: (4) Do you visit homes of friends?, (6) Are you able to go upstairs without using handrails or the wall for support? (10) Do you feel anxious about falling when you walk?, and (25) (In the past two weeks) Have you felt exhausted for no apparent reason?). This short version of KCL can be used in clinical settings for the prediction of frailty status with reasonable accuracy.

## Abbreviations

KCL: Kihon CheckList;

## Data Availability

All data produced in the present study are available upon reasonable request to the authors

## Acknowledgment Author contribution

**Conceptualization:** Motohiko Miyachi, Michihiro Araki, **Data Collection:** Daiki Watanabe, Nobuo Nishi, Takashi Nakagata, Tsukasa Yoshida, Motohiko Miyachi, **Data Analysis:** Masaki Yamamoto, Attayeb Mohsen, **Writing –Original draft:** Attayeb Mohsen, **Writing –Review and editing:** Attayeb Mohsen, Tsukasa Yoshida, Agustin Martin Morales, Michihiro Araki, Motohiko Miyachi, All authors checked the final draft and approved the submission.

